# Weak humoral immune reactivity among residents of long-term care facilities following one dose of the BNT162b2 mRNA COVID-19 vaccine

**DOI:** 10.1101/2021.03.17.21253773

**Authors:** Mark A. Brockman, Francis Mwimanzi, Yurou Sang, Kurtis Ng, Olga Agafitei, Siobhan Ennis, Hope Lapointe, Landon Young, Gisele Umviligihozo, Laura Burns, Chanson Brumme, Victor Leung, Julio S.G. Montaner, Daniel Holmes, Mari DeMarco, Janet Simons, Masa Niikura, Ralph Pantophlet, Marc G. Romney, Zabrina L. Brumme

## Abstract

**Background:** Several Canadian provinces are extending the interval between COVID-19 vaccine doses to increase population vaccine coverage more rapidly. However, immunogenicity of these vaccines after one dose is incompletely characterized, particularly among the elderly, who are at greatest risk of severe COVID-19.

**Methods:** We assessed SARS-CoV-2 humoral responses pre-vaccine and one month following the first dose of BNT162b2 mRNA vaccine, in 12 COVID-19 seronegative residents of long-term care facilities (median age, 82 years), 18 seronegative healthcare workers (HCW; median age, 36 years) and 4 convalescent HCW. Total antibody responses to SARS-CoV-2 nucleocapsid (N) and spike protein receptor binding domain (S/RBD) were assessed using commercial immunoassays. We quantified IgG and IgM responses to S/RBD and determined the ability of antibodies to block S/RBD binding to ACE2 receptor using ELISA. Neutralizing antibody activity was also assessed using pseudovirus and live SARS-CoV-2.

**Results:** After one vaccine dose, binding antibodies against S/RBD were ∼4-fold lower in residents compared to HCW (p<0.001). Inhibition of ACE2 binding was 3-fold lower in residents compared to HCW (p=0.01) and pseudovirus neutralizing activity was 2-fold lower (p=0.003).While six (33%) seronegative HCW neutralized live SARS-CoV-2, only one (8%) resident did (p=0.19). In contrast, convalescent HCW displayed 7- to 20-fold higher levels of binding antibodies and substantial ability to neutralize live virus after one dose.

**Interpretation:** Extending the interval between COVID-19 vaccine doses may pose a risk to the elderly due to lower vaccine immunogenicity in this group. We recommend that second doses not be delayed in elderly individuals.

## INTRODUCTION

Older age is the greatest risk factor for severe COVID-19 outcomes following infection with SARS-CoV-2 ^1-3^. While the rapid development of safe and effective COVID-19 vaccines offers new hope to end this pandemic ^4,5^, the magnitude and durability of vaccine-induced immunity remains unclear, particularly among adults over the age of 80 years who were under-represented in clinical trials ^6,7^ and recent “real world” assessments of vaccine effectiveness ^8-10^. Of four COVID-19 vaccines currently approved for use in Canada, two mRNA vaccines were shown in clinical trials to be highly efficacious when administered as a two-dose regimen, with doses separated by three or four weeks ^11,12^. However, limited availability has prompted many Canadian provinces to delay administration of the second dose of these mRNA vaccines in favor of maximizing the number of individuals benefiting from one dose. This strategy was recently supported by Canada’s National Advisory Committee on Immunization (NACI) ^13^ and is also being employed in the United Kingdom (UK) ^14^. While delayed vaccine dosing is supported by evidence from population-level studies that indicate reductions in the frequency and severity of COVID-19 infections following the initial vaccine rollout, including in LTC facilities, recent COVID-19 outbreaks in LTC facilities underscore the continuing vulnerability of elderly adults to SARS-CoV-2 infection, even after vaccination ^15,16^. Detailed immunogenicity studies following one vaccine dose could help inform public health decision-making for elderly adults living in LTC facilities and the broader community, but few such studies have been conducted, and none to date in Canada.

To address this knowledge gap, we compared the humoral (i.e. antibody-mediated) immune response induced by the first dose of the COVID-19 mRNA vaccine BNT162b2 among elderly residents of LTC facilities compared to younger healthcare workers. We examined serum and plasma collected prior to vaccination, and at approximately 30 days following the first vaccine dose, for the presence of binding antibodies against recombinant SARS-CoV-2 spike (S) protein receptor binding domain (S/RBD). We also quantified antibodies capable of blocking the interaction between S/RBD and the viral entry receptor Angiotensin-Converting Enzyme 2 (ACE2) as well as virus neutralizing antibodies (nAbs).

## METHODS

### Study design and setting

We conducted a prospective cohort study in Vancouver, British Columbia to examine SARS-CoV-2 specific humoral immune responses following one dose of COVID-19 mRNA vaccine among residents of long-term care and assisted living facilities (LTC) and healthcare workers (HCW).

### Ethics approval

Written informed consent was obtained from all participants. This study was approved by the University of British Columbia/Providence Health Care and Simon Fraser University Research Ethics Boards (protocol H20-03906).

### Participants and specimens

Participants were recruited from facilities operated by Providence Health Care (Vancouver, BC) prior to receiving vaccine. Convalescent individuals were identified at study entry based on the presence of serum antibodies recognizing SARS-CoV-2 nucleocapsid (N) using an immunoassay, as described below. Three groups were included in this study: (1) Residents of LTC and assisted living facilities, all seronegative at baseline (n=12); (2) Baseline seronegative HCW (n=18); and (3) Baseline convalescent/seropositive participants, all of whom were HCW (n=4) (**Table 1**). Residents were predominantly male (58%) and a median 82 years of age, while seronegative HCW were predominantly female (72%) and a median of 36 years of age. Convalescent HCW were 50% female and a median 44 years of age. All participants received the BNT162b2 mRNA COVID-19 vaccine developed by Pfizer/BioNTech. Serum and plasma were collected prior to vaccination (23 Dec 2020 – 26 Jan 2021) and again approximately 30 days following the first vaccine dose (18 Jan – 26 Feb 2021). Specimens were processed on the day of collection and frozen until analysis.

**Table 1.** Description of Study Participants

|  | Long-Term Care (LTC) Residents | Healthcare Workers (HCW) | Convalescent (CON) | P-value (LTC vs HCW) |
| --- | --- | --- | --- | --- |
| N | 12 | 18 | 4 | - |
| Female (%) | 5 (42%) | 13 (72%) | 2 (50%) | 0.14 <sup>a</sup> |
| Age, median in yrs [IQR] | 82 [75-87] | 36 [31-50] | 44 [36-48] | <0.0001 <sup>b</sup> |
| Sample Collection, median days post-vaccine [IQR] | 30 [28-30] | 30 [28-31] | 31 [30-33] | 0.11 <sup>b</sup> |

<sup>a</sup> Fisher's Exact Test; <sup>b</sup> Mann-Whitney Test; IQR, interquartile range

### Data sources

Demographic data (age, sex) and COVID-19 vaccination status was collected via participant surveys and confirmed through review of medical records. Antibody immunity data were collected using commercial serological tests and novel immunoassays, as described below.

### Outcome measures

We assessed vaccine-induced antibody responses against SARS-CoV-2 using three measures: (1) Serology assays to detect binding antibodies; (2) ACE2 competition assays to detect receptor-blocking antibodies; and (3) Neutralization assays to detect antibodies that prevent virus infection of cells.

Total binding antibodies in serum against the SARS-CoV-2 nucleocapsid (N) and spike protein receptor binding domain (S/RBD) were determined using the Roche Elecsys Anti-SARS-CoV-2 and Anti-SARS-CoV-2 S assays, respectively, on a cobas e 601 module analyzer (Roche Diagnostics). The former assay provides a qualitative measure of N-specific antibodies that are generated following infection, while the latter assay provides a semi-quantitative measure of total antibodies to S/RBD that are generated following infection or vaccination. Both assays are based on a sandwich ELISA format and utilize electro-chemiluminescence for detection with results reported in arbitrary units/mL, calibrated against an external standard. In addition, we examined the IgG and IgM sub-components of the plasma antibody response to S/RBD using multi-plex bead ELISA on a Luminex 200 instrument. For the assay, recombinant His-tagged S/RBD (USA isolate PC00101P) was cloned into pSELECT-GFPzeo (InvivoGen) and expressed by transfection of 293T cells (ATCC) using Lipofectamine 2000 reagent (Invitrogen). S/RBD was purified by chromatography (TALON Superflow, Millipore Sigma) and coupled to carboxylated xMAP beads (Bio-Rad). Plasma samples were diluted 1:200 in phosphate buffered saline prior to incubation with beads. Bound IgG or IgM was detected using PE-conjugated secondary antibodies (BioLegend) and results are reported as mean fluorescence intensities (MFI).

The ability of plasma antibodies to block ACE-2 receptor was also determined by Luminex ELISA, however with addition of a soluble recombinant ACE2 protein at a saturating concentration (>1 *µ*M final concentration; Abcam) during incubation of plasma with S/RBD beads. We calculated the difference in bound IgG in the absence of ACE2 versus the presence of ACE2 and report the results as MFI.

NAbs were determined using pseudovirus and live SARS-CoV-2 infectivity assays employing VeroE6-TMPRSS2 cells (JCRB-1818) as targets. Pseudotyped virions were generated by co-transfecting 293T cells (ATCC) with a plasmid encoding SARS-CoV-2 Wuhan-Hu-1 Spike glycoprotein (Genscript) or VSV-G Indiana glycoprotein control (pHEF-VSV-G; NIH HIV Reagent Program) along with a lentiviral packaging plasmid (pCMV-dR8.2 dvpr ^17^ ; Addgene) and luciferase expression plasmid (pLenti CMV Puro LUC (w168-1) ^18^ ; Addgene). Pseudovirus-containing supernatant was harvested on day 3 and used immediately. For neutralization assays, virus-containing supernatant was added to diluted plasma in DMEM supplemented with 10% FBS, 20 *µ*g/mL polybrene and Pen/Strep, incubated for 1 h at 37°C and then added to target cells pre-seeded in 96-well plates in duplicate. Luciferase activity was assayed 3 days thereafter and percentage of neutralizing activity was calculated relative to cell-only and virus-only controls. Results are reported as relative luciferase units (RLU). Live virus neutralization assays were performed using SARS-CoV-2 isolate USA-WA1/2020 (BEI Resources) in a Containment Level 3 facility. Viral stock was adjusted to 50 TCID_50_/200 *µ*l in DMEM in the presence of serial 2-fold dilutions of plasma, incubated for 1 hour at 4°C and then added to target cells pre-seeded in 96-well plates in triplicate. The appearance of viral cytopathic effect (CPE) was recorded 3 days post-infection. Neutralizing activity is reported as present if CPE was prevented in at least one of three wells following incubation with 1:20 diluted plasma or is reported as the highest reciprocal dilution where CPE was prevented in all wells.

### Statistical analysis

For most measures, we applied the Mann-Whitney U-test to compare rank values between residents and HCW. Neutralization frequencies in live-virus neutralization assays were examined using the Fisher’s exact test. All tests were two-tailed, with an alpha of 0.05 considered statistically significant. Analyses were conducted using Prism v9.0.2 (GraphPad).

## RESULTS

### Humoral immunity to one dose of BNT162b2 mRNA vaccine

Using a range of immunoassays, we observed significantly lower antibody responses to SARS-CoV-2 following one dose of BNT162b2 mRNA vaccine among 12 residents of LTC facilities compared to 18 HCW (**Figure** and **Table 2**). In contrast, and consistent with prior studies ^19,20^, highly elevated antibody responses were seen in four convalescent HCW after one dose of vaccine.

**Figure.**
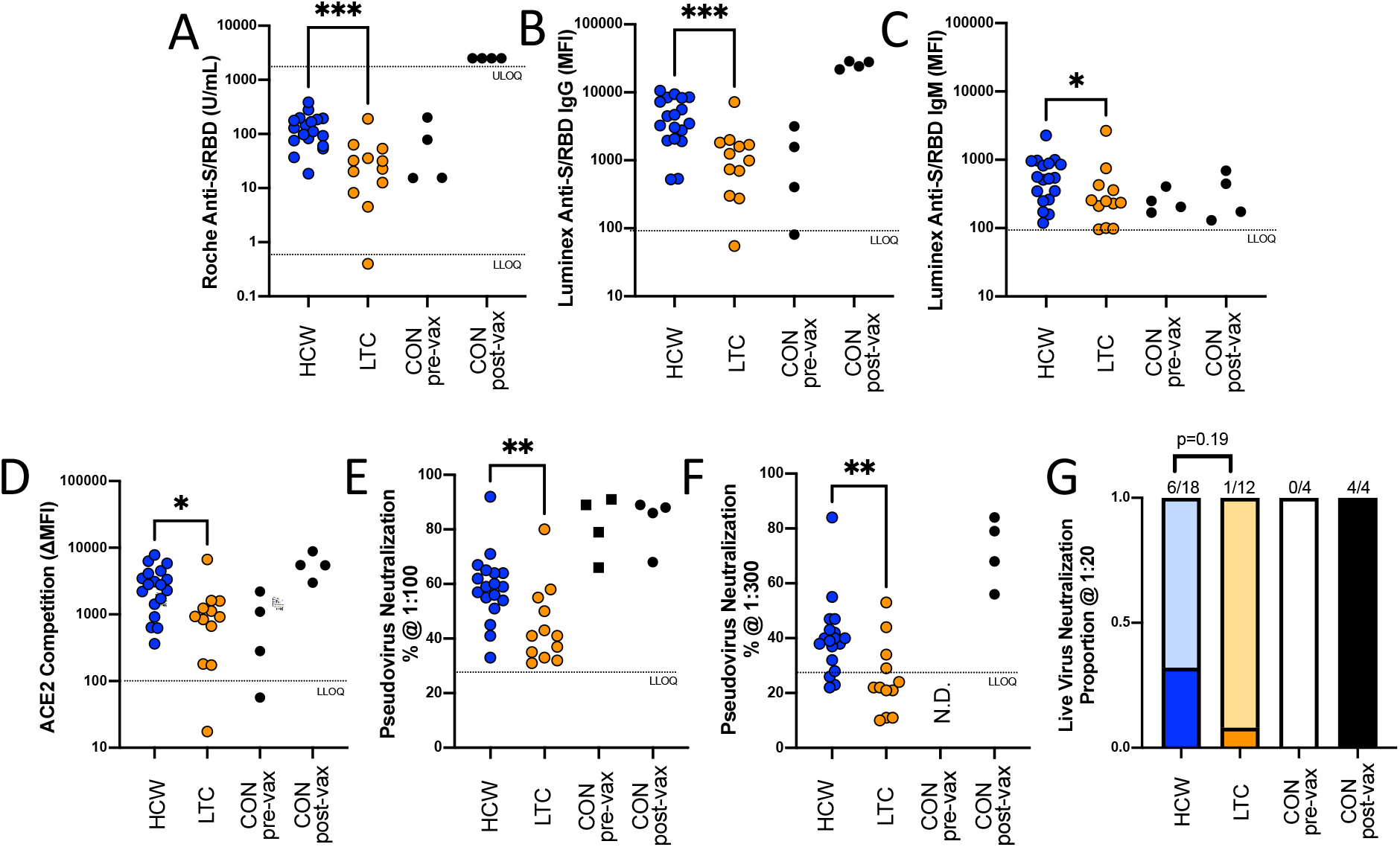
Humoral immune responses to the first dose of BNT162b2 are lower in long-term care residents compared to healthcare workers. (A, B, C) Binding antibodies to the spike protein receptor binding domain (S/RBD) were measured in serum and plasma following one dose of vaccine. Results are displayed for (A) total binding antibodies in sera, in Units/ml (Roche Elecsys), (B) IgG in 1:200 diluted plasma, as mean fluorescence intensity (MFI) (Luminex), and (C) IgM in 1:200 diluted plasma, as MFI (Luminex). (D) Results of ACE2 competition assays are displayed as the change in MFI following coincubation of plasma with soluble ACE2 (Luminex). (E, F) Results of pseudovirus neutralization assays are shown as the percent reduction in luminescence signal using plasma dilutions of 1:100 (E) or 1:300 (F). (G) Results of live SARS-CoV-2 neutralization assays are shown as the proportion of participants that display neutralizing activity using a plasma dilution of 1:20. Where available, results for convalescent participants shown at baseline (prior to vaccination; pre-vax) and after one dose of vaccine (post-vax). Abbreviations: HCW, healthcare worker (n=18); LTC, long-term care resident (n=12); CON, convalescent (n=4); LLOQ, lower limit of quantification; ULOQ, upper limit of quantification. Statistical analyses (A-F) were conducted using the Mann-Whitney U-test: *, p<0.05; **, p<0.005; ***, p<0.0005. Statistical analysis in G was conducted using the Fisher’s Exact test.

**Table 2.** Humoral immune responses in study participants

| Assay Description | Measure | Long-Term Care (LTC) Residents (n=12) |  | Healthcare Workers (HCW) (n=18) |  | Convalescent (n=4) |  | P-value<br>(LTC vs HCW, post vaccine) |
| --- | --- | --- | --- | --- | --- | --- | --- | --- |
|  |  | Baseline (pre-vaccine) | Post-vaccine | Baseline (pre-vaccine) | Post-vaccine | Baseline (pre-vaccine) | Post-vaccine |  |
| Serum_anti-N (Roche) | Units/mL | <0.4 | <0.4 | <0.4 | <0.4 | 44 [14-83] | 33 [12-71] | N/A |
| Serum_anti-S/RBD (Roche) | Units/mL | <0.4 | 27 [9-49] | <0.4 | 120 [72-188] | 47 [15-171] | >2500 | 0.0002 <sup>a</sup> |
| Plasma_anti-S/RBD_IgG (Luminex) | MFI | <80 | 1123 [401-1795] | <80 | 3966 [2041-8327] | 986 [162-2761] | 26013 [22364-28495] | 0.0005 <sup>a</sup> |
| Plasma_anti-S/RBD_IgM (Luminex) | MFI | <80 | 242 [128-410] | <80 | 538 [259-905] | 228 [178-367] | 310 141-630] | 0.43 <sup>a</sup> |
| ACE2 Competition (Luminex) | MFI | <80 | 927 [302-1506] | <80 | 2799 [1294-4224] | N.D. | 5479 [3605-8005] | 0.012 <sup>a</sup> |
| Pseudovirus neutralization (1:100) | % Inhibition | <30 | 41 [34-54] | <30 | 59 [53-64] | 84 [69-91] | 87 [73-89] | 0.035 <sup>a</sup> |
| Pseudovirus neutralization (1:300) | % Inhibition | <30 | <30 [<30-33] | <30 | 40 [31-44] | N.D. | 74 [59-83] | 0.0041 <sup>a</sup> |
| SARS-CoV-2 neutralization (1:20) | # Positive (%) | 0 (0%) | 1 (8%) | 0 (0%) | 6 (33%) | 0 (0%) | 4 (100%) | 0.19 <sup>b</sup> |
| SARS-CoV-2 neutralization (Titer) | Reciprocal Dilution | <20 | <20 | <20 | <20 (<20-20) | <20 | 240 [100-560] | 0.16 <sup>a</sup> |

Values are reported as medians and interquartile ranges; <sup>a</sup> Mann-Whitney U-test; <sup>b</sup> Fisher's Exact Test; <, below assay quantification limit; > above assay quantification limit; N.D., not determined

First, we examined sera and plasma for the presence of binding antibodies to recombinant SARS-C0V-2 S/RBD. After one vaccine dose, median serum binding antibodies against S/RBD were 4.4-fold lower in samples from residents compared to those from HCW using the Elecsys SARS-CoV-2 S assay (p=0.0002), with no measurable response detected for one resident sample (**Figure, A**). For context, median serum binding antibodies to S/RBD in residents, collected at peak following one vaccine dose, were 1.7-fold lower than those from convalescent participants at study entry (pre-vaccine) who were estimated to be on average 7 months post-infection (**Figure, A**). Similarly, following one vaccine dose, median plasma IgG antibodies to S/RBD were 3.5-fold lower in residents compared to HCW using ELISA (p=0.0005), with the same resident exhibiting no response above background (**Figure, B**). For context, median plasma IgG antibodies to S/RBD in residents at peak following a single vaccine dose were comparable to those of convalescent participants at study entry (**Figure, B**). After one dose of vaccine, serum binding antibodies to S/RBD increased >50-fold and plasma IgG antibodies increased >25-fold among the convalescent participants (**Figure, A, B**), consistent with “boosting” of pre-existing immunity in this group. Therefore, total and IgG binding antibodies achieved by one vaccine dose in convalescent participants were >90- and >20-fold higher respectively than those in residents (and >20- and 6.5-fold higher than those in previously seronegative HCW). Residents also displayed 2.2-fold lower median plasma IgM binding antibodies compared to HCW following vaccination (p=0.04) (**Figure, C**); however, these values did not differ substantially from convalescent participants either before or after vaccination.

Next, we conducted competition ELISA assays to measure the potential ability of plasma antibodies to block critical interactions between S/RBD and its cellular receptor ACE2. We found that residents displayed 3-fold lower ACE2 blocking activity compared to HCW after one dose of vaccine (p=0.01) (**Figure, D**). In contrast, ACE2 blocking activity in vaccinated HCW was similar to that achieved in convalescent participants after one vaccine dose (**Figure, D**).

Lastly, we performed viral neutralization assays to more directly assess the ability of plasma antibodies to block infection of target cells, which may involve epitopes on SARS-CoV-2 Spike located outside of S/RBD ^21^. Using a pseudovirus assay, we observed that samples from residents displayed 1.5- to 2-fold lower median neutralizing activities compared to HCW after one vaccine dose (p<0.005 at 1:100 and 1:300 plasma dilution) (**Figure, E, F**). Notably, this response in HCW was itself 1.5- to 1.9-fold lower than that observed in convalescent participants after vaccination (p<0.005 at 1:100 and 1:300 dilution). Using a more stringent live SARS-CoV-2 assay, we observed that plasma from six (33%) HCW displayed modest evidence of neutralizing activity following one dose of vaccine, whereas plasma from only one (8%) resident did so (p=0.19 at 1:20 dilution). For comparison, plasma from all four convalescent participants displayed substantial neutralizing activity against live SARS-CoV-2 following one vaccine dose (median reciprocal titer of 240), despite plasma from these individuals showing no neutralizing activity prior to vaccination.

## INTERPRETATION

### Summary

We examined antibody responses to SARS-CoV-2 induced by one dose of BNT162b2 mRNA COVID-19 vaccine among 12 seronegative residents of LTC facilities, 18 seronegative healthcare workers (HCW) and four convalescent (baseline seropositive) participants using a combination of commercial and research immunoassays. Overall, we observed significantly weaker binding and nAb responses among elderly residents compared to HCW. Notably, vaccine-induced antibody responses in HCW were significantly lower than those in convalescent participants, indicating that a single dose of BNT162b2 is not sufficient to induce robust nAbs, even in younger persons.

### Explanation

The magnitude and durability of immune responses to COVID-19 vaccines remain incompletely characterized ^22^, particularly among the elderly after only the first of a two-dose mRNA vaccine regime. Our observation of lower humoral responses to the BNT162b2 mRNA vaccine among residents of LTC facilities is nevertheless consistent with prior literature describing poor reactivity to vaccines in older adults that often necessitates the use of higher vaccine antigen concentrations, stronger adjuvants and additional booster shots ^23-25^. To our knowledge, this is only the second study to report on the strength of humoral immunity in elderly participants following one dose of mRNA vaccine, and the first in Canada. The other report from researchers in the UK, which is available as a pre-print ^26^, demonstrated similar age-related impairments in binding and neutralizing antibodies following one dose of BNT162b2 in 42 participants, including 18 over the age of 80 years, while T cell responses were similar between younger and older participants. The authors of that study also showed that nAbs remained lower among participants over 80 years of age even after receiving a second vaccine dose.

Taken together with the UK study results, our observations of significantly weaker antibody responses in elderly individuals following one dose of mRNA vaccine are highly relevant in the context of recent recommendations by Canada’s NACI to extend the interval between first and second COVID-19 mRNA vaccines doses to up to 121 days in all persons. Indeed, recent outbreaks in LTC facilities in British Columbia and Ontario underscore the ongoing vulnerability of elderly adults to SARS-CoV-2 even after vaccination. The finding that one elderly resident in our study (of 12 in total, **Figure**) failed to generate a detectable binding antibody response after one vaccine dose further illustrates this risk in at least a subset of individuals. Critically, it remains unclear if the observed decrease in the frequency of COVID-19 outbreaks in LTC facilities following the initial vaccine rollout is primarily attributable to individual-level immunity achieved in the elderly residents or whether it can be explained by localized herd immunity, as most HCW and essential staff were vaccinated at the same time as residents. If protection is largely due to the latter, then the decision to delay second doses may pose more substantial hazards to seniors living in the community, who, unlike LTC residents, will remain at elevated risk of exposure to infection by the largely unvaccinated population that surrounds them. LTC residents whose second doses are now delayed may also be at elevated risk to unvaccinated HCW and members of the community as LTC facilities accept more visitors.

### Future directions

This report reflects an interim analysis of 34 participants for whom specimens were available from study entry and at approximately 30 days following the first vaccine dose, with the goal of rapidly disseminating observations that may inform decision-making on extended vaccine dose intervals in Canada. We continue to characterize immune responses following one and two doses of vaccine in our larger cohort.

### Limitations

Since immune correlates of protection for SARS-CoV-2 transmission and disease severity are unknown, the implications of our results for individual- and population-level immunity from COVID-19 are presently unclear. Indeed, the level of nAbs induced in the majority of LTC residents following one vaccine dose may be sufficient to prevent infection (or severe disease) in at least some of the individuals. Our sample size was small; despite this, the weaker antibody responses observed among LTC residents were statistically significant for almost all measures of humoral immunity tested here and our results are highly consistent with those from a recent UK study. Participant sex distribution differed between groups and though not statistically significant (p=0.16, **Table 1**) may have modulated study outcomes. Due to limited sample size, we are unable to determine the independent effects of sex and age in our data; however, we observed significantly lower serum binding antibodies in residents compared to HCW after restricting our analysis to only female participants (p=0.02, data not shown), suggesting that age is a greater determining factor. With the exception of the Roche Elecsys assays, we relied on novel immunoassays developed by our team that have not been formally externally validated. Nevertheless, and despite the use of different target antigens and anticipated differences in assay stringency, the high concordance observed among various methods strengthens confidence in our observations. Finally, we have not yet assessed the impact of a second vaccine dose on humoral immunity in our cohort, as the earliest participants received their first doses in late December 2020/January 2021 only. We thus prioritized this interim analysis in light of its potential immediate relevance to decision-making related to COVID-19 vaccine dosing intervals in Canada.

### Conclusion

Extending the interval between BNT162b2 mRNA COVID-19 vaccine doses may put the elderly disproportionately at risk due to lesser vaccine immunogenicity in this group. Incomplete vaccine-induced immunity after on dose may be particularly risky for elderly individuals living in the community, whose main contacts will remain largely unvaccinated for some time. We recommend that second doses of the COVID-19 mRNA vaccines not be delayed in the elderly, the segment of society that has already borne the largest brunt of the COVID-19 pandemic.

## Data Availability

All study data is available to qualified investigators by request to the corresponding authors. A data transfer agreement may be required.

## ACKNOWLEDGEMENTS

We thank the leadership and staff of Providence Health Care, including long-term care and assisted living residences, for their support of this study, and laboratory staff at St. Paul’s Hospital, the BC Centre for Excellence in HIV/AIDS and Simon Fraser University for assistance. We also acknowledge Haisheng Yu for plasmid 2019-nCov_pcDNA3.1(+)-P2A-eGFP (Genscript # MC_0101087), Lung-Ji Chang for plasmid pHEF-VSV-G (HIV Reagent Program # ARP-4693), Bob Weinberg for pCMV-dR8.2 dvpr (Addgene plasmid # 8455), and Eric Campeau & Paul Kaufman for pLenti CMV Puro LUC (w168-1) (Addgene plasmid # 17477). Above all, we thank the participants, without whom this study would not be possible.

